# Mental health outcomes among front and second line health workers associated with the COVID-19 pandemic in Italy

**DOI:** 10.1101/2020.04.16.20067801

**Authors:** Rodolfo Rossi, Valentina Socci, Francesca Pacitti, Giorgio Di Lorenzo, Antinisca Di Marco, Alberto Siracusano, Alessandro Rossi

**Affiliations:** Chair of Psychiatry, Department of Systems Medicine, University of Rome Tor Vergata, Rome, Italy; Chair of Psychiatry, Department of Applied Clinical Sciences and Biotechnologies, University of L’Aquila, L’Aquila, Italy; Department of information engineering, computer science and mathematics, University of L’Aquila, L’Aquila, Italy

## Abstract

In this study, we report on mental health outcomes among health workers (HWs) involved with the COVID-19 pandemic in Italy.

Data on mental health on 1379 HWs were collected between March 27^th^ and March 31^th^ 2020 using an on-line questionnaire spread throughout social networks, using a snowball technique along with sponsored social network advertisement. Key mental health outcomes were Post-Traumatic Stress Disorder symptoms (PTSD), severe depression, anxiety, insomnia and perceived stress.

PTSD symptoms, severe depression, anxiety and insomnia, and high perceived stress were endorsed respectively by 681 (49.38%), 341 (24.73%), 273 (19.80%), 114 (8.27%) and 302 (21.90%) respondents. Regression analysis show that younger age, female gender, being a front-line HWs, having a colleague deceased, hospitalised or in quarantine were associated with poor mental health outcomes.

This is the first report on mental health outcomes and associated risk factors among HWs associated with the COVID-19 pandemic in Italy, confirming a substantial proportion of health workers involved with the COVID-19 pandemic having mental health issues, in particular young women, first-line HWs.

## Introduction

It has been recently shown that health workers (HWs) involved with the COVID-19 pandemic in Whuan and the Hubei province are exposed to high levels of stressful or traumatic events and express substantial negative mental health outcomes^1^, including stress-related symptoms, depression, anxiety and insomnia.

To the best of our knowledge, no data on HWs involved with the COVID-19 pandemic in Europe are available so far.

In this study, we report on mental health outcomes, using an on-line survey, among HWs in Italy.

## Methods

Cross-sectional web-based observational study. Data on mental health were collected between March 27^th^ and March 31^th^ 2020 using an on-line questionnaire spread throughout social networks, using a snowball technique along with sponsored social network advertisement. According to Italian public health organizations, the sampling period corresponded to the days immediately preceding the COVID-19 contagion peak, associated with a high level of healthcare system overwhelming. All HWs reporting to work in Italy were eligible. The questionnaire investigated key demographic variables, workplace characteristics, such as being a front-line or second-line worker, and information regarding the direct impact of the COVID-19, including having colleagues infected or deceased. Key mental health outcomes were Post-Traumatic Stress Disorder symptoms (PTSD), severe depression, anxiety, insomnia and perceived stress. These were assessed using the Italian version of the Global Psychotrauma Screen (GPS)^2^, the 9-item Patient Health Questionnaire (PHQ-9)^3^, the 7-item Generalized Anxiety Disorder scale (GAD-7)^4^, the 7-item Insomnia Severity Index (ISI)^5^, and the 10-item Perceived Stress Scale (PSS)^6^. Participants were classified as endorsing the aforementioned condition according to the following cut-offs: suspect PTSD ≥3 on the GPS-PTSD subscale, severe depression if PHQ≥15, severe anxiety if GAD ≥15, severe insomnia if ISI≥22. Because the PSS does not represent any official disorder, a quartile split was used to separate the first quartile from the remaining participants.

A seemingly unrelated multivariable logistic regression model was fitted in order to explore the impact of gender, age, first-line working position, occupation, self and colleagues’ exposure to contagion on the selected outcomes. Seemingly unrelated regression models allow to jointly model correlated outcomes.

Approval for this study was obtained from the local IRB at University of L’Aquila. On-line consent was obtained from the participants.

## Results

A total of 1379 HWs completed the questionnaire. Because of the web-based snowball sampling strategy, response rate could not be calculated. Sample characteristics are reported in table 1. PTSD symptoms, severe depression, anxiety and insomnia, and high perceived stress were endorsed respectively by 681 (49.38%), 341 (24.73%), 273 (19.80%), 114 (8.27%) and 302 (21.90%) respondents (Table 1).

**Table 1:** Sample characteristics. North: Aosta Valley, Piedmont, Liguria, Lombardy, Trentino-Alto Adige, Veneto, Friuli-Venezia Giulia, Emilia Romagna; Center: Tuscany, Marche, Umbria, Lazio; South: Abruzzo, Molise, Campania, Apulia, Basilicata, Calabria, Sicily, Sardinia. GP: General Practitioner; HCA: Health Care Assistant; GPS-PTSD: Global Psychotrauma Scale - post traumatic stress disorder subscale; PHQ: Patient Health Questionnaire; GAD: Generalized anxiety disorder scale; ISI: Insomnia Severity Index; PSS: Perceived Stress Scale; IQR: Interquartile range.

|  | **Overall** | **North** | **Center** | **South** |
| --- | --- | --- | --- | --- |
|  | *No. (%) / Median (IQR)* | *No. (%) / Median (IQR)* | *No. (%) / Median (IQR)* | *No. (%) / Median (IQR)* |
| **No.** | 1379 | 667/1359 (48.3%) | 412/1359 (29.8%) | 280/1359 (20.3%) |
| **Gender** |  |  |  |  |
| *Women* | 1064/1379 (77.2%) | 533/667 (79.9%) | 321/412 (77.9%) | 194/280 (69.3%) |
| *Men* | 315/1379 (22.8%) | 134/667 (20.1) | 91/412 (22.1%) | 86/280 (30.7%) |
| **Age** |  |  |  |  |
|  | 39.0 (16.0) | 38.0 (16.0) | (41.0) (16.0) | 38.0 (15.0) |
| **Working Position** |  |  |  |  |
| *Front-line* | 725 (52.57%) | 448/667 (67.17%) | 181/412 (43.93%) | 88/279 (31.43%) |
| *Second-line* | 653 (47.35%) | 219/667 (32.83%) | 231/412 (56.07%) | 191/279 (68.21%) |
| **Occupation** |  |  |  |  |
| *Nurse* | 472/1378 (34.23%) | 265/667 (39.7%) | 131/412 (31.8%) | 67/279 (23.93%) |
| *Physician* | 433/1378 (31.40%) | 163/667 (24.44%) | 164/412 (39.81%) | 100/279 (35.71%) |
| *GP* | 86/1378 (6.24%) | 35/667 (5.25%) | 22/412 (5.34%) | 28/279 (10.00%) |
| *Other* | 275/1378 (19.94%) | 137/667 (20.54%) | 77/412 (18.69%) | 58/279 (20.71%) |
| *HCA* | 112/1378 (8.12%) | 67/667 (10.04%) | 18/412 (4.37%) | 26/279 (9.29%) |
| **Education Level** |  |  |  |  |
| *Undergraduate* | 222/1373 (16.10%) | 131/667 (19.64%) | 49/410 (11.95%) | 36/276 (13.04%) |
| *Postgraduate* | 1151/1373 (83.47%) | 536/667 (80.36%) | 361/410 (88.05%) | 240/276 (86.96%) |
| **GPS-PTSD$\geq 3$** |  |  |  |  |
|  | 681/1376 (49.38%) | 352 (52.77%) | 193 (46.84%) | 127/280 (45.85%) |
| **PHQ $\geq 15$** |  |  |  |  |
|  | 341/1378 (24.73%) | 182/666 (27.29%) | 99/412 (24.03%) | 55/280 (19.64%) |
| **GAD $\geq 15$** |  |  |  |  |
|  | 273/1378 (19.80%) | 130/666 (19.49%) | 84/412 (20.39%) | 55/280 (19.64%) |
| **ISI $\geq 22$** |  |  |  |  |
|  | 114/1378 (8.27%) | 66/667 (9.90%) | 28/412 (6.80%) | 17/280 (6.07%) |
| **PSS 1<sup>st</sup> quartile** |  |  |  |  |
|  | 302/1378 (21.90%) | 160/667 (23.99%) | 86/412 (20.87%) | 52/280 (18.57%) |
| **GPS total score** |  |  |  |  |
|  | 9 (6) | 9 (6) | 9 (6) | 8 (7) |
| **PHQ total score** |  |  |  |  |
|  | 10 (9) | 10 (9) | 10 (9) | 8 (8) |
| **GAD total score** |  |  |  |  |
|  | 9 (9) | 9 (8) | 9 (10) | 8 (9.5) |
| **ISI total score** |  |  |  |  |
|  | 10 (12) | 11 (12) | 10 (10.5) | 8 (13) |
| **PSS total score** |  |  |  |  |
|  | 24 (11) | 24 (10) | 24 (9) | 22 (12) |

18 participants were excluded from regression analysis due to missing data. Regression analysis show that younger age and female gender were associated with all of the investigated outcomes, except for insomnia. Being a front-line HWs was specifically associated with PTSD symptoms. General Practitioners were more likely to endorse PTSD symptoms, nurses and health care assistants were more likely to endorse severe insomnia. Having a colleague deceased, hospitalized or in quarantine was associated with PTSD symptoms, depression, insomnia and perceived stress. Being exposed to contagion was associated with depression (Table 2).

**Table 2:** Seemingly Unrelated Logistic Regression Analysis. GP: General Practitioner; HCA: Health Care Assistant; GPS-PTSD: Global Psychotrauma Scale - post traumatic stress disorder subscale; PHQ: Patient Health Questionnaire; GAD: Generalized anxiety disorder scale; ISI: Insomnia Severity Index; PSS: Perceived Stress Scale. *p<0.05; **p<0.005; ***p<0.001. OR: odds ratio. CI: Confidence Intervals.

| N=1369 | GPS - PTSD |  | PHQ |  | GAD |  | ISI |  | PSS |  |
| --- | --- | --- | --- | --- | --- | --- | --- | --- | --- | --- |
|  | OR | 95% CI | OR | 95% CI | OR | 95% CI | OR | 95% CI | OR | 95% CI |
| **Age std** | 0.69** | [0.53,0.88] | 0.74* | [0.56,0.98] | 0.60** | [0.44,0.82] | 0.71 | [0.46,1.09] | 0.63** | [0.46,0.85] |
| **Gender** |  |  |  |  |  |  |  |  |  |  |
| Male | 1.00 (ref) | [1.00,1.00] | - |  | - |  | - |  | - |  |
| Female | 2.31*** | [1.76,3.05] | 2.03*** | [1.44,2.87] | 2.18*** | [1.49,3.19] | 1.38 | [0.82,2.33] | 2.64*** | [1.80,3.87] |
| **Working Position** |  |  |  |  |  |  |  |  |  |  |
| Second-line | 1.00 (ref) | [1.00,1.00] | - |  | - |  | - |  | - |  |
| First-line | 1.37* | [1.05,1.80] | 1.04 | [0.76,1.42] | 1.13 | [0.80,1.59] | 1.27 | [0.78,2.07] | 1.16 | [0.84,1.60] |
| **Occupation** |  |  |  |  |  |  |  |  |  |  |
| Other HW | 1.00 (ref) | [1.00,1.00] | - |  | - |  | - |  | - |  |
| Nurse | 1.12 | [0.81,1.55] | 1.36 | [0.95,1.96] | 1.09 | [0.74,1.61] | 2.03* | [1.14,3.59] | 0.74 | [0.51,1.08] |
| Physician | 1.20 | [0.86,1.67] | 0.71 | [0.48,1.05] | 0.96 | [0.64,1.44] | 0.89 | [0.46,1.72] | 0.75 | [0.51,1.11] |
| GP | 1.75* | [1.03,2.97] | 0.98 | [0.53,1.82] | 1.05 | [0.53,2.08] | 1.47 | [0.56,3.87] | 1.18 | [0.66,2.11] |
| HCA | 0.95 | [0.60,1.52] | 1.18 | [0.70,1.98] | 1.05 | [0.60,1.84] | 2.34* | [1.06,5.18] | 0.59 | [0.33,1.05] |
| **Colleagues affected** |  |  |  |  |  |  |  |  |  |  |
| No affected colleagues | 1.00 (ref) | [1.00,1.00] | - |  | - |  | - |  | - |  |
| Deceased colleague | 2.60** | [1.30,5.19] | 2.07* | [1.05,4.07] | 0.97 | [0.41,2.29] | 2.94* | [1.21,7.18] | 1.84 | [0.88,3.87] |
| Infected and hospitalized | 1.54* | [1.10,2.16] | 1.39 | [0.95,2.03] | 1.18 | [0.78,1.77] | 1.14 | [0.66,1.96] | 1.93** | [1.30,2.85] |
| Infected and in quarantine | 1.59*** | [1.21,2.09] | 1.38 | [1.00,1.90] | 1.19 | [0.85,1.67] | 0.88 | [0.54,1.45] | 1.66** | [1.19,2.32] |
| **Not exposed to contagion** | 1.00 (ref) | [1.00,1.00] |  |  |  |  |  |  |  |  |
| Exposed to contagion | 1.23 | [0.93,1.62] | 1.54* | [1.11,2.14] | 1.14 | [0.81,1.62] | 1.45 | [0.88,2.39] | 1.01 | [0.73,1.41] |

## Discussion

This is the first report on mental health outcomes and associated risk factors among HWs associated with the COVID-19 pandemic in Italy. These results are in line with prior reports from China, confirming a substantial proportion of health workers involved with the COVID-19 pandemic having mental health issues, in particular young women, first-line HWs. Our results warrant further monitoring and specific interventions on health workers throughout the COVID-19 pandemic in order to prevent long-term mental-health related disabilities.

## Data Availability

data available upon request from the authors

## Acknowledgments

This work is supported by Territori Aperti, a project founded by “Fondo Territori Lavoro e Conoscenza CGIL CISL UIL”.

